# Social Determinants of Health Data Quality at Different Levels of Geographic Detail

**DOI:** 10.1101/2023.02.16.23286036

**Authors:** Melody L. Greer, Maryam Y. Garza, Steve Sample, Sudeepa Bhattacharyya

**Author notes:** Corresponding Author: Melody L. Greer,.

## Abstract

Social determinants of health (SDOH) impact 80% of health outcomes from acute to chronic disorders, and attempts are underway to provide these data elements to clinicians. It is, however, difficult to collect SDOH data through (1) surveys, which provide inconsistent and incomplete data, or (2) aggregates at the neighborhood level. Data from these sources is not sufficiently accurate, complete, and up-to-date. To demonstrate this, we have compared the Area Deprivation Index (ADI) to purchased commercial consumer data at the individual-household level. The ADI is composed of income, education, employment, and housing quality information. Although this index does a good job of representing populations, it is not adequate to describe individuals, especially in a healthcare context. Aggregate measures are, by definition, not sufficiently granular to describe each individual within the population they represent and may result in biased or imprecise data when simply assigned to the individual. Moreover, this problem is generalizable to any community-level element, not just ADI, in so far as they are an aggregate of the individual community members.

## 1. Introduction

Social determinants of health (SDOH) impact 80% of health outcomes [1] from acute to chronic disorders. Interest in this information has been increasing over the last decade and the Centers for Medicare & Medicaid Services (CMS) Comprehensive Primary Care Plus (CPC+) Model requires providers to assess patients’ social risks to aid in more accurate care delivery [2]. Attempts are underway to consistently provide these data elements to researchers and clinicians. It is, however, difficult to collect SDOH data as the main mechanisms used are (1) surveys which require time for documentation, and are not standardized across institutions, [3] or (2) aggregates at the neighborhood level [4]. In order to provide value, SDOH data must be complete, current, accurate. But neighborhood level data elements are not specific to each individual and so may not be correct for every patient. To study this relationship and understand the level of error that results from using neighborhood level data as a stand-in for individual level data we compared the Area Deprivation Index to individual and their household level social determinants.

The ADI is built from census data and includes the factors of income, education, employment, and housing quality [5]. The American Community Survey five-year estimates are used to produce the ADI which is a neighborhood level index available freely for download at the state or national level in both census blocks and ZIP Code versions. This validated index has been available for 30 years and has seen increased usage over the last several years. The ADI does a good job of representing populations [6]. It does not, however, purport to represent individuals and should therefore not be used as a replacement for individual level representations. Aggregate measures do not describe each individual within the population they represent. For example, if certain SDOH elements like education or income are highly variable within a neighborhood, the neighborhood-level aggregate will be an imprecise estimate of any individual’s actual measure. This idea is generalizable to any community-level data element as they are an aggregate of the measures of individual community members.

## 2. Methods

We created an integrated data set composed of approximately 55,000 electronic health records linked to the state level ADI and commercial consumer data. The ADI was obtained by downloading the census block group level ADI score and the zip+4 centroids contained within each block group. In previous work we have developed a repeatable SDOH enrichment and integration process to incorporate dynamically evolving SDOH domain concepts from consumers into clinical data. [7] In the course of that work we demonstrated that commercial consumer data can be a viable source of SDOH factors at an individual-level for clinical data providing a path for clinicians to improve patient treatment and care. [8, 9]

Because ADI is composed of income, education, employment, and housing quality data we have chosen these elements from the commercial consumer data for comparison. We computed the average ADI within each zip code. Commercial consumer data elements were not available for every individual or their household in the study, and those with null values were eliminated from the calculations, but were retained for other data quality comparisons.

## 3. Results

As shown in Figures 1-4 the box plots reflect the variance of the summarized ADI within the different levels of each demographic element and demonstrates the potential for inaccuracy when applying neighborhood level data to an individual. Although the neighborhood level elements skew as expected the overlap in actual values with respect to disparity is problematic. This overlap is a result of the wide variance for each of the categories.

**Figure 1.**
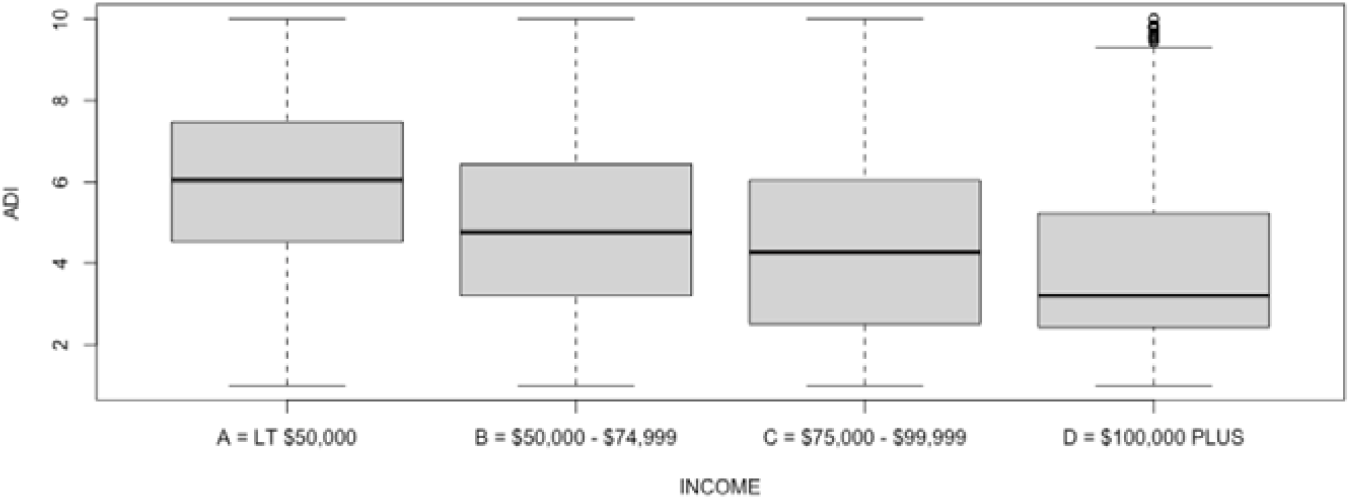
Income value compared with ADI.

**Figure 2.**
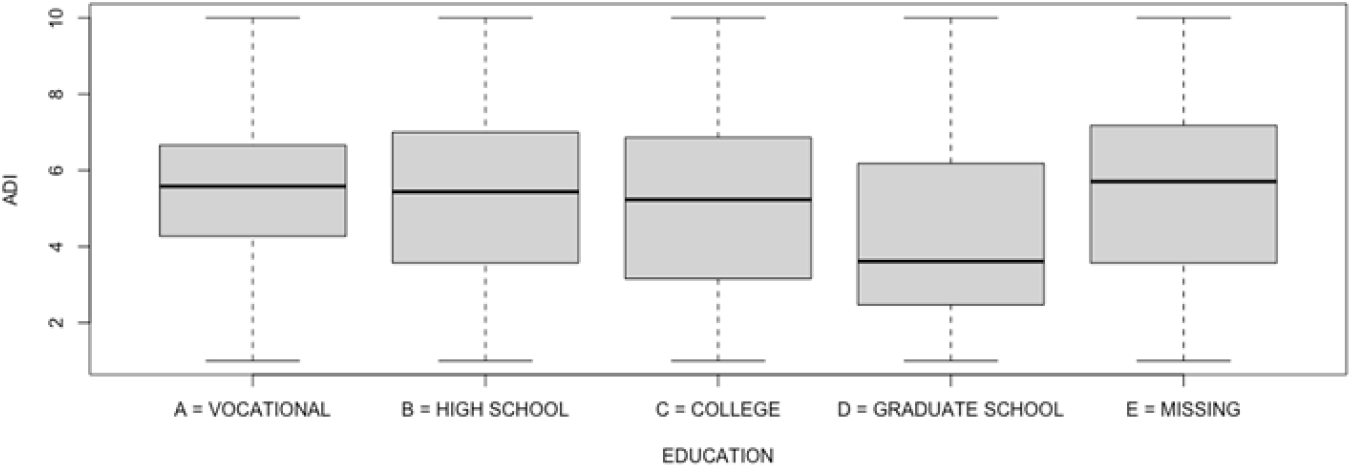
Education level compared with ADI.

In addition to accuracy, it is important to consider other aspects of data quality for use in patient care, such as completeness and timeliness. Upon review of both datasets, commercial consumer data vs. ADI, there were clear differences with regards to data completion. Typically, commercial consumer data is available for many individuals, but may not be complete across all factors or data elements (e.g., for people who use only cash, have no subscriptions, and do not use discount cards). The commercial consumer data we used in this analysis was 98.98% complete for income, 62.63% complete for education, 48.23% complete for employment, and 94.69% complete for housing quality across all individuals in the dataset. In comparison, because the ADI is an aggregate measure, data for all factors was ‘complete’ for every individual or their household.

In regards to timeliness, the commercial consumer data was no more than six months old when purchased whereas ADI, and other indices constructed using census data, can only be as current as the last five-year estimate. Because commercial consumer data is purchased for an agreed-upon amount of time that can include a regular refresh schedule, it is always current.

## 4. Discussion

This work demonstrates that neighborhood level SDOH elements are not sufficiently granular to provide clinicians or researchers individual level information. Other researchers have conducted related research with different deprivation indices and have come to similar conclusions. [4] Further, research suggests that the relationship between health outcomes and neighborhood level social determinants of health are not caused by the neighborhood environment but result from sorting by economic means [10].

### 4.1. Accuracy

The box plots reflect the range of ADI values within each category, revealing the broad range of deprivation for households and individuals. This demonstrates that ADI is associated on a large geographic scale but not on a smaller one. The standard deviation and variance of income, education, employment, and housing quality shown in Figures 1-4 are clear indications that these population level elements would often be inaccurate if used for individual patients in medical decision-making. Doing so opens the door for misclassification of patient risk level and invalidates the output of any would be automated clinical decision support systems.

**Figure 3.**
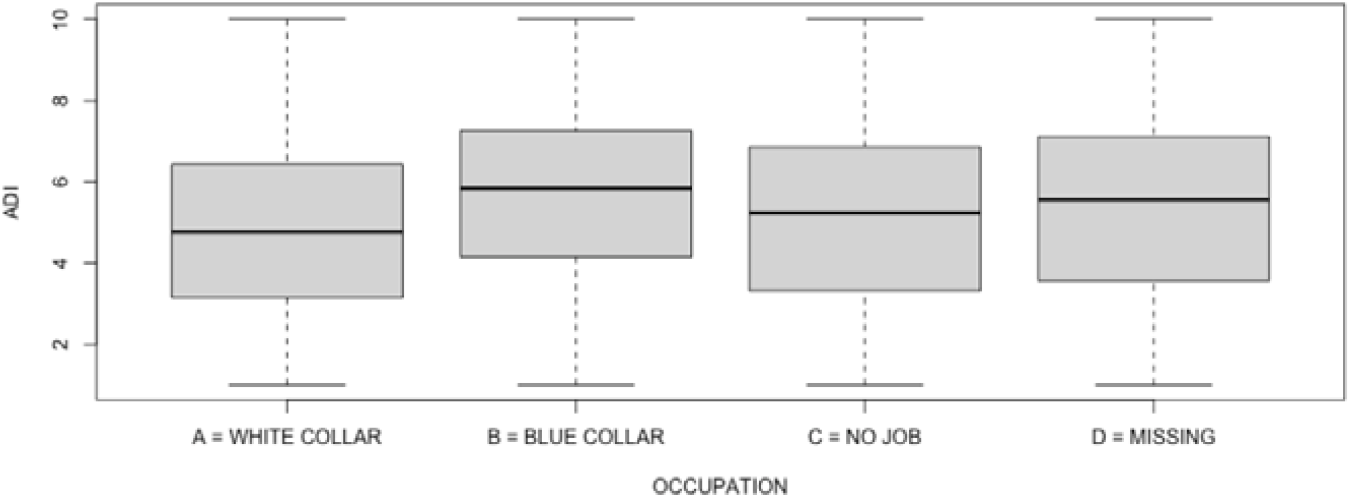
Occupation value compared with ADI.

**Figure 4.**
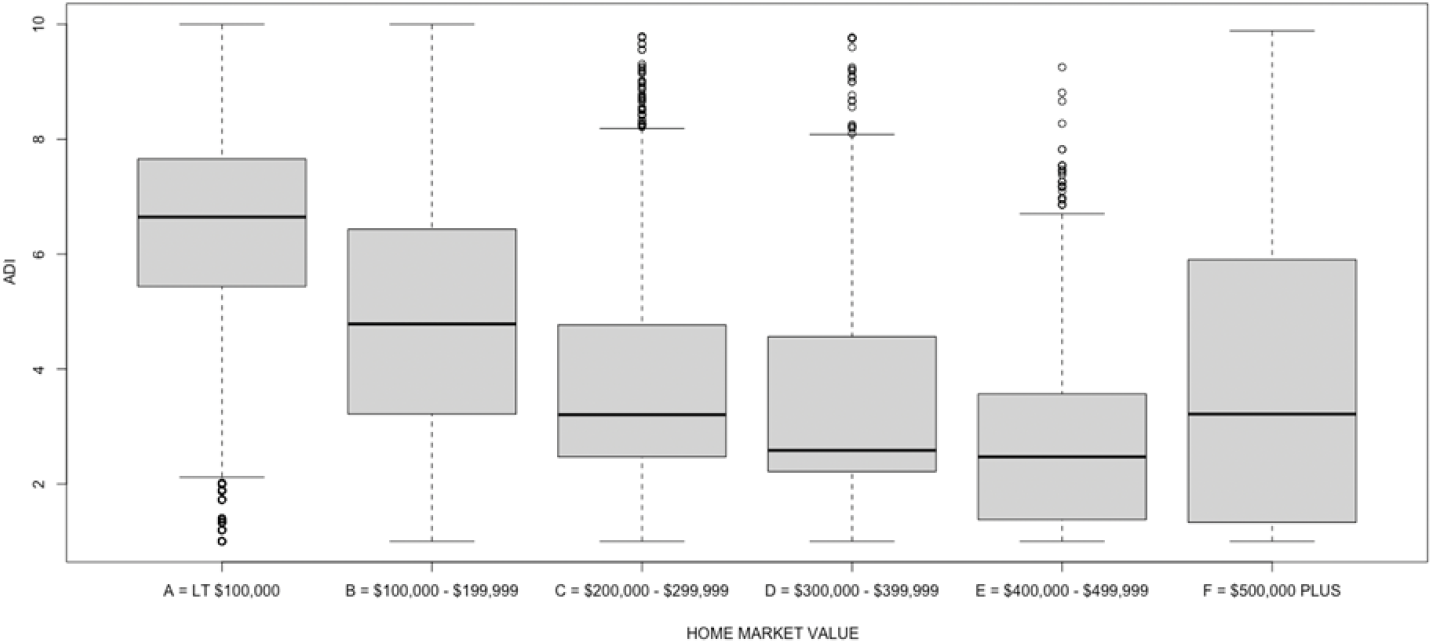
Home market value compared with ADI.

### 4.2. Completeness

Missing information can affect the quality of care a patient receives. This problem is not as serious as wrong information, but can still have a negative impact on medical decision-making, especially if the problem is large and pervasive. Because ADI is an aggregate value, completeness is not an issue. However, a weakness of commercial consumer data is that it is not always available for every individual or their household. In this work, we have ignored null values in an effort to focus on accuracy, but this cannot be a solution for clinical applications.

### 4.3. Timeliness

For healthcare purposes, it is important to have current social context information. Frequently, individuals will change habits, move, and switch employers. These changes directly impact factors related to their health in both positive and negative ways. If the information provided to clinicians or clinical decision support systems is out of date, this could have a negative impact on patients by delaying access to care. Commercial consumer data can be constantly refreshed to maintain the optimal current status. As far as we are aware, this is not true for publicly available data, and it is definitely not the case for any elements that use census data.

## 5. Conclusions

Because social context information is valuable in clinical practice and research, SDOH data elements have received an increasing amount of attention in the last two decades. Consistently collecting this data in a standard format remains a problem. Clinical providers are extremely busy, and additional data entry is not an optimal solution. In addition, SDOH screening instruments are not standardized, making interoperability challenging, if not impossible. To address this difficulty, social data aggregated at the neighborhood level has been proposed as a surrogate. However, this solution is also problematic as we have seen with the ADI because individual level data is not equivalent to neighborhood level data in every case. Moreover, aggregate measures are, by definition, not on target for each individual within the population which they represent. As a result, we conclude that data used to understand the social context for healthcare should be complete, current, accurate, and specific to the individual and their unique household.

## Data Availability

Data are private and are not available

